# Seroprevalence and Trends of Transfusion-Transmissible Infections Among Blood Donors in the Volta Region, Ghana: A Four-Year Retrospective Study

**DOI:** 10.64898/2026.05.04.26352407

**Authors:** Shadrack Ayanku, Lois Akuba, Caroline Tetteh, Timothy Yao Akweh, Eyram Kuma Hanu, David Annor Kwasie, Stephen Bekpenaneso Bawu, Klutse Fianko, Anthony Zunuo Dongdem

## Abstract

**Background:** Blood transfusion is a life-saving intervention; however, transfusion-transmissible infections (TTIs) such as human immunodeficiency virus (HIV), hepatitis B virus (HBV), hepatitis C virus (HCV), and syphilis remain major public health concerns, particularly in low-and middle-income countries. This study assessed the seroprevalence and temporal trends of TTIs among blood donors in the Volta Region of Ghana and to identify the demographic factors associated with seropositivity

**Methods:** A retrospective cross-sectional study was conducted using secondary data from blood donors at Ho Teaching Hospital and Hohoe Regional Hospital between January 2020 and December 2023. Data from 6,147 eligible donors were extracted and analyzed using STATA version 17. Descriptive statistics summarized prevalence, while chi-square or Fisher’s exact tests assessed associations. Multivariable logistic regression was used to identify predictors of TTI seropositivity at a 5% significance level.

**Results:** The overall prevalence of TTIs was 8.1%, with syphilis (3.6%) being the most prevalent infection, followed by HBV (1.8%), HCV (1.8%), and HIV (1.0%). All infections peaked in 2022 before declining in 2023. Older age (≥50 years) and year of donation were significant predictors of TTI positivity. In Hohoe, male donors had lower odds of HCV infection compared to females (aOR = 0.13; 95% CI: 0.06–0.28).

**Conclusions:** Although TTI prevalence was relatively low, temporal increases and age-related disparities highlight the need for strengthened donor screening, targeted recruitment of voluntary donors, and enhanced surveillance strategies to ensure blood safety.

## Introduction

Blood transfusion is a life-saving medical intervention that has been in use for decades. However, the transmission of transfusion-transmissible infections (TTIs) through blood transfusion remains a significant public health concern (1). Transfusion transmissible infections, such as HIV, hepatitis B and C, and syphilis, can be transmitted through blood transfusions (2).

Globally, there are approximately 170 million individuals chronically infected with hepatitis C virus (HCV), more than 350 million with hepatitis B virus (HBV), and 38 million human immunodeficiency virus (HIV)-infected people (3).

In Sub-Saharan Africa, the overall prevalence of TTIs was reported in Ethiopia at 4.1%, Eritrea at 3.6%, Malawi at 10.7%, Eastern Democratic Republic of Congo (D.R.C) at 14.7%, Western Kenya at 9.4%, Northern Tanzania at 10.1%, whereas in Southwestern Uganda, the burden of TTIs was estimated at 5.67% (4).

In Ghana, similar to many other low- and middle-income countries (LMICs), the prevalence of TTIs among blood donors is a significant concern. The overall TTI prevalence is 21.0%, with specific rates for HBV (6.6%), HCV (4.9%), HIV (2.9%), and Syphilis (6.8%) (5).

A study conducted in Ashaiman municipality reported the prevalence of different TTIs as follows: HBV (4.9%), HCV (1.5%), and Syphilis (4.0%) (1). A study conducted in Akatsi South Municipality in the Volta region of Ghana also reported an 8.0% TTI prevalence, including HIV (3.8%), HBV (3.2%), and HCV (1.0%) (6).

The World Health Organization (WHO) recommends screening all donated blood for TTIs to ensure the safety of the blood supply (7). Despite these recommendations, the prevalence of TTIs among blood donors in Ghana remains high, with studies reporting varying prevalence rates depending on the region and population studied (1). This highlights the need for ongoing surveillance, particularly in areas like Hohoe and Ho, to better understand and mitigate the risks associated with TTIs in blood donation. This study therefore aimed to determine the seroprevalence and temporal trends of transfusion-transmissible infections (TTIs) among blood donors in Hohoe and Ho, Volta Region, Ghana, and to identify demographic factors associated with seropositivity.

## Methods

### Study Design

A retrospective cross-sectional study design was employed using secondary data from blood bank records. This design was appropriate for estimating the prevalence of TTIs and examining associations between donor characteristics and TTI seropositivity over the study period

### Study Site Description

The study was conducted at the Ho Teaching Hospital Blood Bank and the Hohoe Regional Hospital Blood Bank, both located in the Volta Region of Ghana. Ho Teaching Hospital is a tertiary referral facility and a major training center for the University of Health and Allied Sciences, serving the southern Volta Region and neighboring areas. Hohoe Regional Hospital is a key referral facility for the northern Volta and Oti Regions. Both blood banks collect blood from voluntary and family replacement donors and routinely screen all donated blood for transfusion-transmissible infections (TTIs) including HIV, hepatitis B virus (HBV), hepatitis C virus (HCV), and syphilis, following national blood safety guidelines

### Study population

The study population included all individuals who donated blood at the two facilities between January 2020 and December 2023.

### Inclusion Criteria

All blood donors whose records contained complete demographic information and TTI screening results for HIV, HBV, HCV, and Syphilis between January 2020 and December 2023 were included in the study.

### Exclusion Criteria

Donor records that were incomplete, missing key demographic details, or lacking any of the four TTI test results were excluded from the study.

### Variables

The primary outcome variable was TTI seropositivity (HIV, HBV, HCV, and syphilis). Independent variables included age, sex, and year of donation.

### Sample Size Determination

All available donor records that met the eligibility criteria during the study period were included. A total of 6,147 donor records were reviewed, comprising 2,015 donors from Ho Teaching Hospital and 4,132 donors from Hohoe Regional Hospital. The inclusion of all eligible records ensured adequate statistical power to estimate prevalence and assess associations.

### Sampling Method

The Ho Teaching Hospital and the Volta Regional Hospital were purposively selected for this study due to their roles as major referral centers in the Volta Region and their availability of comprehensive and reliable blood donor screening data. These facilities maintain structured records on transfusion-transmissible infections (TTIs), making them suitable for retrospective analysis.

### Data Collection

Blood donors were screened using standard blood bank procedures involving donor interview, physical examination, and laboratory testing. During the interview, key information such as age, sex, medical history, previous donations, medication use, recent illnesses, travel history, and risk behaviors is asked. Physical assessment including; weight, blood pressure, pulse rate, temperature, and hemoglobin level are measured. Blood samples are then collected from eligible donors and screened for transfusion-transmissible infections including HIV, Hepatitis B, Hepatitis C, and syphilis using Rapid Diagnostic Tests (RDTs) and ELISA. Only donors tested negative to these pathogens are accepted for donation.

### Data Extraction

Data was extracted from blood bank registers from May to June 2025, using a structured data extraction form developed for the study. Variables collected included donor age, sex, year of donation, and serological screening results for HIV, HBV, HCV, and syphilis. To ensure data quality, extracted records were cross-checked for completeness and consistency before analysis. Personal identifiers were not collected.

### Statistical Analysis

Data was entered into Microsoft Excel and exported to STATA version 17 for analysis. Descriptive statistics (frequencies, percentages, means, and standard deviations) were used to summarize donor characteristics and TTI prevalence. Associations between demographic variables and TTI status were assessed using chi-square or Fisher’s exact tests, as appropriate. Multivariable logistic regression models were fitted to identify independent predictors of TTI seropositivity, and adjusted odds ratios (aORs) with 95% confidence intervals (CIs) were reported. Statistical significance was set at p < 0.05.

## Results

### Demographic Characteristics

A total of 6,147 blood donors were included in the analysis, comprising 2,015 (32.8%) from Ho and 4,132 (67.2%) from Hohoe. The overall mean age of donors was 26.9 ± 7.4 years, with donors in Ho being slightly older (28.2 ± 7.9 years) than those in Hohoe (26.3 ± 7.1 years). Majority of donors were aged 20–29 years (55.6%), followed by 30–39 years (23.5%). Donors aged ≥50 years constitute less than 1% of the study population. The donor population was predominantly male (95.1%), with females accounting for only 4.9%

### Overall Prevalence of Transfusion-Transmissible Infections

The overall prevalence of transfusion-transmissible infections (TTIs) among blood donors was low. The prevalence of HIV was 1.0% (63/6,147), HBV 1.8% (113/6,147), HCV 1.8% (109/6,147), and syphilis 3.6% (219/6,147). Among the TTIs, syphilis was the most prevalent infection, followed by HBV and HCV, which showed similar prevalence levels.

Trends of Transfusion-Transmissible Infections (2020–2023)

Figure 1 shows a clear temporal variation in TTI prevalence over the study period. Across all infections, prevalence was relatively low in 2020 and 2021, followed by a marked increase in 2022, and a subsequent decline in 2023.

**Figure 1.**
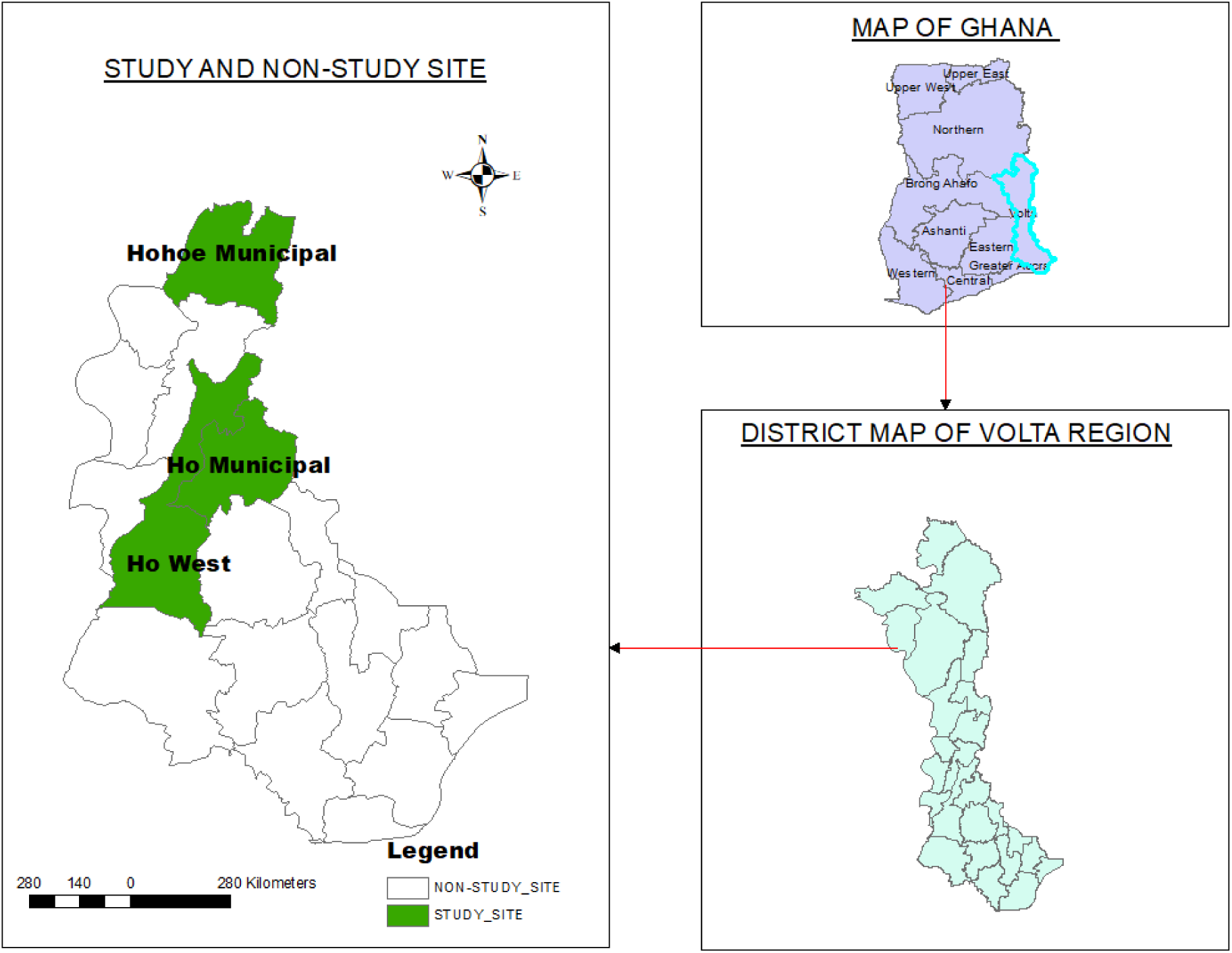
showing study and non-study site.

**Figure 2.**
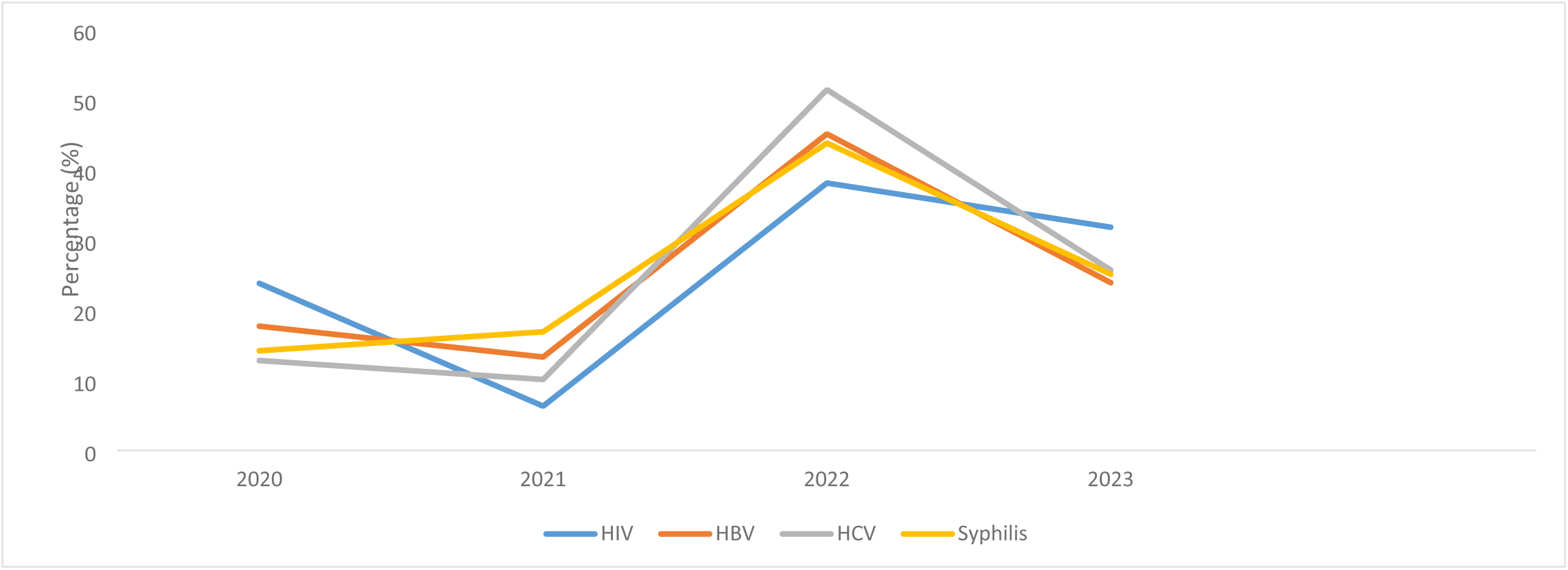
Trends of Transfusion-Transmissible Infections.

Specifically, HIV prevalence increased from 0.3% in 2021 to 1.6% in 2022, while HBV rose from 0.9% to 3.5%, HCV from 0.7% to 3.8%, and syphilis from 2.3% to 6.6% during the same period. Although prevalence declined in 2023, it remained higher than levels observed in 2020–2021 for most infections.

### Site-Specific Prevalence of TTIs

#### Ho Blood Bank

Among donors in Ho (n = 2,015), the prevalence of HIV, HBV, HCV, and syphilis was 1.3%, 3.7%, 1.8%, and 3.1%, respectively. HBV was the most prevalent infection in this site. A notable increase in HBV (8.5%) and syphilis (6.4%) was observed in 2022, followed by a decline in 2023. (Table 3a)

**Table 1.** Demographic Characteristics of the Blood Donors.

| Variable | Place |  |  |
| --- | --- | --- | --- |
|  | Ho n (%) | Hohoe n (%) |  |
| **Mean age (SD)** | 28.2 (±7.9) | 26.3 (±7.1) | 26.9 (±7.4) |
| **Age group** |  |  |  |
| <20 | 187(9.3) | 624(15.1) | 811 (13.2) |
| 20-29 | 1069(53.1) | 2351(56.9) | 3420 (55.6) |
| 30-39 | 546(27.1) | 901(21.8) | 1447 (23.5) |
| 40-49 | 182(9.0) | 236(5.7) | 418 (6.8) |
| 50-59 | 31(1.5) | 20(0.5) | 51 (0.8) |
| **Sex** |  |  |  |
| Female | 133(6.6) | 171(4.1) | 304 (4.9) |
| Male | 1882(93.4) | 3961(95.9) | 5843 (95.1) |

**Table 2:** Prevalence of transfusion-transmissible infections among blood donors in the blood banks.

| variable | HIV | Prevalence (%) | HBV | Prevalence (%) | HCV | Prevalence (%) | Syphilis | Prevalence (%) | Total screened |
| --- | --- | --- | --- | --- | --- | --- | --- | --- | --- |

| Year | Positive | negative |  | Positive | negative |  | Positive | negative |  | Positive | Negative |  |  |
| --- | --- | --- | --- | --- | --- | --- | --- | --- | --- | --- | --- | --- | --- |
| 2020 | 15 | 1626 | 0.9 | 20 | 1621 | 1.2 | 14 | 1627 | 0.9 | 31 | 1610 | 1.9 | 1641 |
| 2021 | 4 | 1607 | 0.3 | 15 | 1596 | 0.9 | 11 | 1600 | 0.7 | 37 | 1574 | 2.3 | 1611 |
| 2022 | 24 | 1435 | 1.6 | 51 | 1408 | 3.5 | 56 | 1403 | 3.8 | 96 | 1363 | 6.6 | 1459 |
| 2023 | 20 | 1416 | 1.4 | 27 | 1409 | 1.9 | 28 | 1408 | 1.9 | 55 | 1381 | 3.8 | 1436 |
| Total | 63 | 6084 | 1.0 | 113 | 6034 | 1.8 | 109 | 6038 | 1.8 | 219 | 5928 | 3.6 | 6147 |

**Table 3a:** Prevalence of transfusion-transmissible infections (TTIs) among blood donors at Ho blood banks.

| variabl<br>e | HIV |  | Prevale<br>nce (%) | HBV |  | Prevalen<br>ce (%) | HCV |  | Prevalenc<br>e (%) | Syphilis |  | Prevalenc<br>e (%) | Total<br>screene<br>d |
| --- | --- | --- | --- | --- | --- | --- | --- | --- | --- | --- | --- | --- | --- |
| Year | posi<br>tive | negativ<br>e |  | Positiv<br>e | negativ<br>e |  | Positiv<br>e | negativ<br>e |  | Positiv<br>e | negativ<br>e |  |  |
| 2020 | 11 | 533 | 2.0 | 16 | 528 | 2.9 | 9 | 535 | 1.7 | 9 | 535 | 1.7 | 544 |
| 2021 | 3 | 603 | 0.5 | 10 | 596 | 1.7 | 8 | 598 | 1.3 | 11 | 595 | 1.8 | 606 |
| 2022 | 5 | 432 | 1.1 | 37 | 400 | 8.5 | 17 | 420 | 3.9 | 28 | 409 | 6.4 | 437 |
| 2023 | 8 | 420 | 1.9 | 12 | 416 | 2.8 | 3 | 425 | 0.7 | 14 | 414 | 3.3 | 428 |
| Total | 27 | 1988 | 1.3 | 75 | 1940 | 3.7 | 37 | 1978 | 1.8 | 62 | 1953 | 3.1 | 2015 |

#### Hohoe Blood Bank

Among donors in Hohoe (n = 4,132), the prevalence of HIV, HBV, HCV, and syphilis was 0.9%, 0.9%, 1.7%, and 3.8%, respectively. Syphilis was the most prevalent infection in this site. Similar, to Ho, all infections peaked in 2022, particularly HCV (3.8%) and syphilis (6.7%), with a reduction observed in 2023. (Table 3b)

**Table 3b:** Prevalence of transfusion-transmissible infections (TTIs) among blood donors at Hohoe blood banks.

| variable | HIV |  | Prevalence (%) | HBV |  | Prevalence (%) | HCV |  | Prevalence (%) | Syphilis |  | Prevalence (%) | Total screened |
| --- | --- | --- | --- | --- | --- | --- | --- | --- | --- | --- | --- | --- | --- |
| Year | Positive | Negative |  | Positive | negative |  | Positive | negative |  | Positive | negative |  |  |
| 2020 | 4 | 1093 | 0.4 | 4 | 1093 | 0.4 | 5 | 1092 | 0.5 | 22 | 1075 | 2.0 | 1097 |
| 2021 | 1 | 1004 | 0.1 | 5 | 1000 | 0.5 | 3 | 1002 | 0.3 | 26 | 979 | 2.6 | 1005 |
| 2022 | 19 | 1003 | 1.9 | 14 | 1008 | 1.4 | 39 | 983 | 3.8 | 68 | 954 | 6.7 | 1022 |
| 2023 | 12 | 996 | 1.2 | 13 | 995 | 1.3 | 25 | 983 | 2.5 | 41 | 967 | 4.1 | 1008 |
| Total | 36 | 4096 | 0.9 | 36 | 4096 | 0.9 | 72 | 4060 | 1.7 | 157 | 3975 | 3.8 | 4132 |

### Factors Associated with TTIs: Bivariate Analysis

#### Ho Blood Bank

In Ho, age and year of donation were significantly associated with several TTIs. Increasing age was associated with higher prevalence of HIV (p = 0.02), HBV (p = 0.01), and HCV (p = 0.03). Year of donation was significantly associated with HBV (p < 0.01) and syphilis (p < 0.01), with higher prevalence observed in 2022.

No statistically significant association was observed between gender and any of the TTIs.

#### Hohoe Blood Bank

In Hohoe, age and year of donation were also significantly associated with TTIs. Age was significantly associated with HBV (p < 0.01), HCV (p = 0.02), and syphilis (p < 0.01), with higher prevalence observed among older donors.

Gender was significantly associated with HCV infection (p < 0.01), with higher prevalence observed among females compared to males. Year of donation were significantly associated with all TTIs (p < 0.01), with peak prevalence consistently observed in 2022.

**Table 4a:**
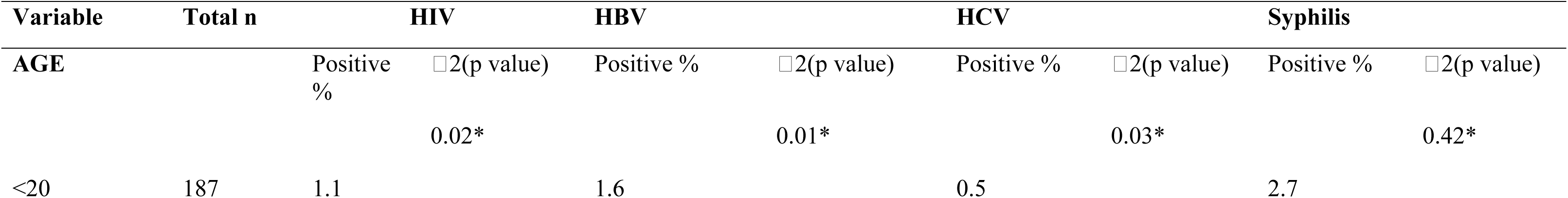
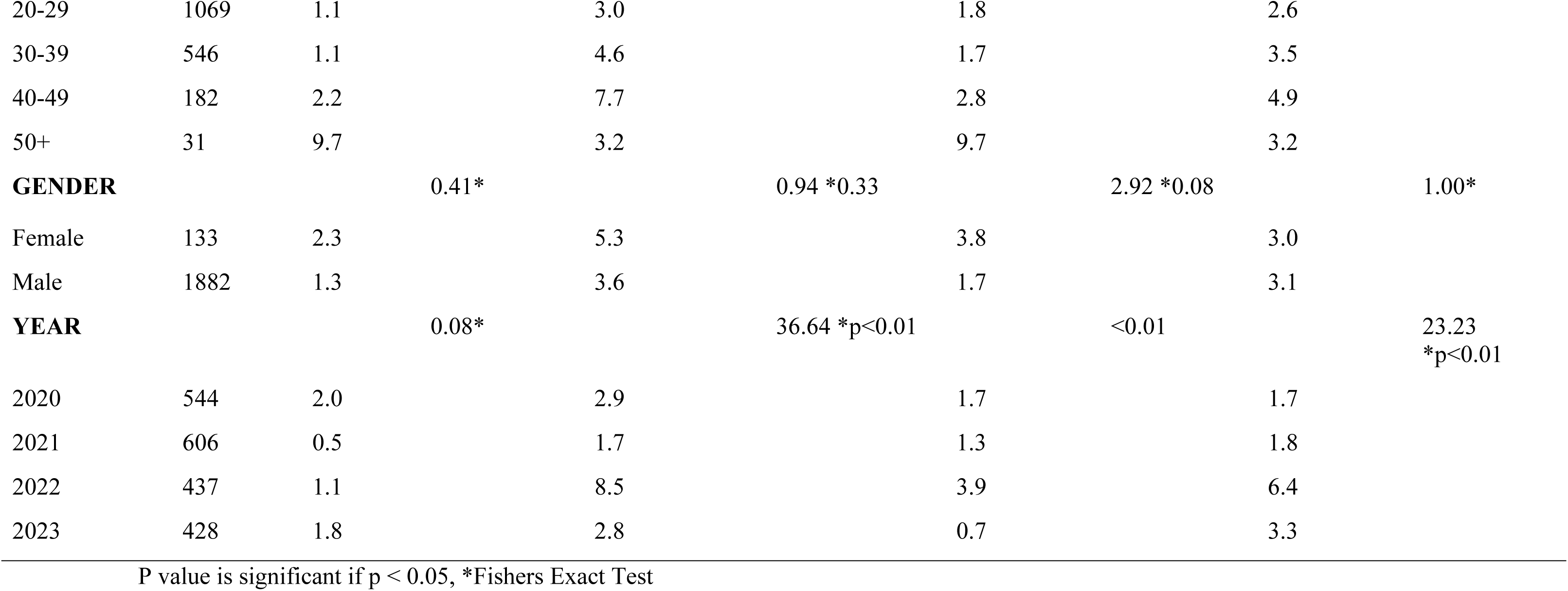
Association Between Demographic Characteristics and Transfusion-Transmissible Infections Among Blood Donors in Ho.

**Table 4b:**
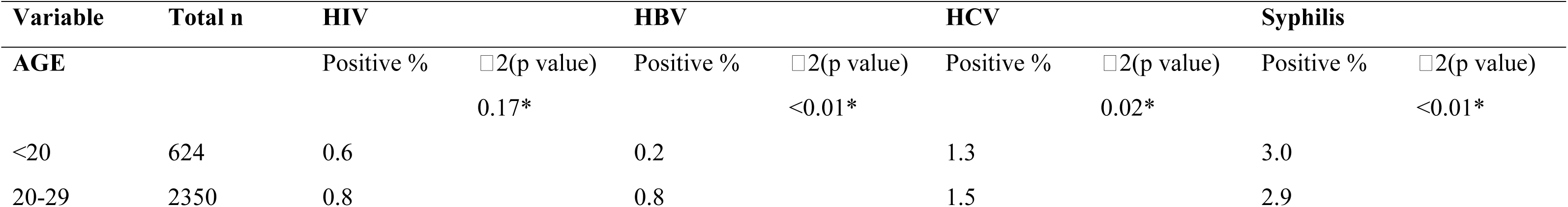
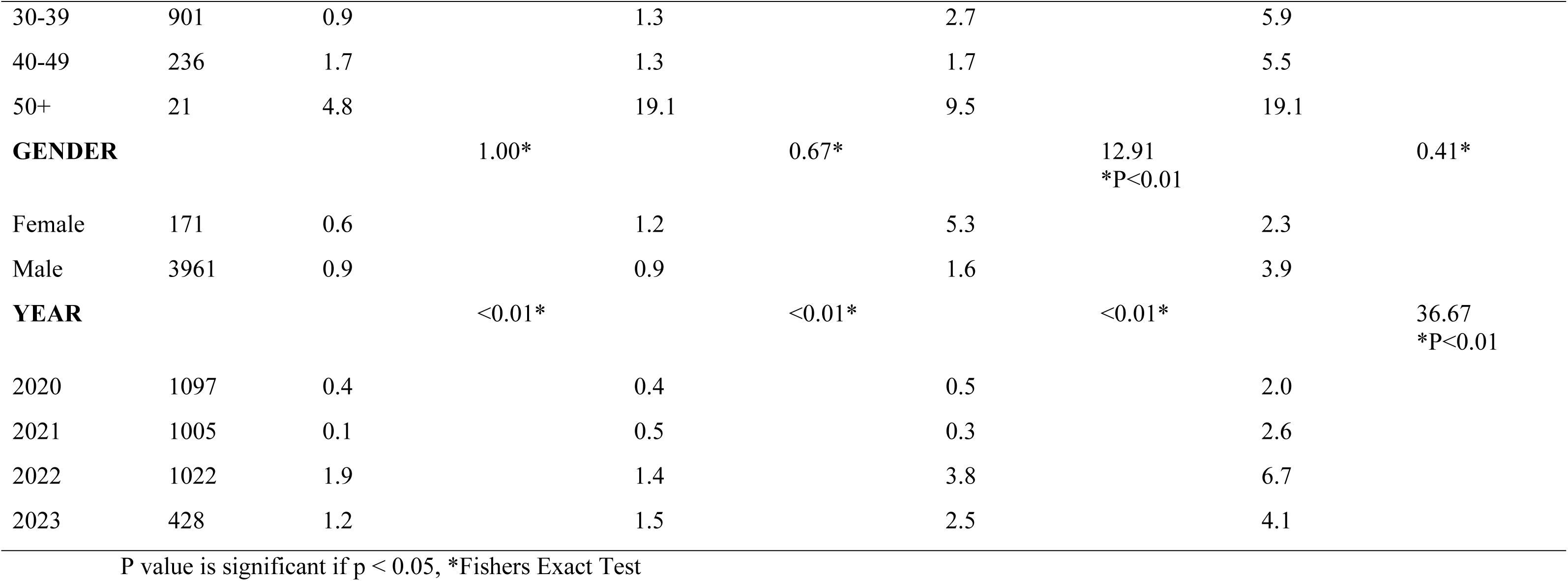
Association Between Demographic Characteristics and Transfusion-Transmissible Infections Among Blood Donors in Hohoe.

#### Ho Blood Bank

After adjusting for potential confounders, age and year of donation remained significant predictors of TTI positivity.

Donors aged ≥50 years had significantly higher odds of HIV infection (aOR = 8.67; 95% CI: 1.37–54.56) and HCV infection (aOR = 21.66; 95% CI: 2.03–230.75) compared to donors aged <20 years.

Donors aged 40–49 years also had increased odds of HBV infection (aOR = 4.36; 95% CI: 1.21–15.61). The year of donation was significantly associated with the incidence of the infections. Compared to 2020, donors in 2022 had higher odds of HBV (aOR = 2.90; 95% CI: 1.58–5.34), HCV (aOR = 2.35; 95% CI: 1.03–5.35), and syphilis (aOR = 3.99; 95% CI: 1.85–8.60).

Gender was not significantly associated with any TTI after adjustment.

#### Hohoe Blood Bank

In Hohoe, age, gender, and year of donation were significant predictors of TTIs.

Donors aged ≥50 years had significantly higher odds of HBV (aOR = 82.16; 95% CI: 7.41–910.79), HCV (aOR = 13.14; 95% CI: 2.38–72.42), and syphilis (aOR = 9.40; 95% CI: 2.58–34.15) compared to donors aged <20 years.

Male donors had significantly lower odds of HCV infection compared to females (aOR = 0.13; 95% CI: 0.06–0.28).

Year of donation were strongly associated with all TTIs. Compared to 2020, donors in 2022 had significantly higher odds of HIV (aOR = 5.43; 95% CI: 1.83–16.09), HBV (aOR = 4.06; 95% CI: 1.26–11.48), HCV (aOR = 11.84; 95% CI: 4.56–30.75), and syphilis (aOR = 3.46; 95% CI: 2.12–5.63). Elevated odds persisted in 2023 for all infections.

**Table 5a:** Multivariable logistic regression analysis of factors associated with transfusion-transmissible infections among Ho blood donors.

| Variable | HIV | HBV | HCV | Syphilis |
| --- | --- | --- | --- | --- |
| **AGE** | aOR (95% CI) | aOR (95% CI) | aOR (95% CI) | aOR (95% CI) |
| <20 | Ref | Ref | Ref | Ref |
| 20-29 | 1.01 (0.22, 4.65) | 1.72 (0.51, 5.72) | 3.31 (0.43, 25.29) | 0.85 (0.32, 2.26) |
| 30-39 | 0.96 (0.18, 4.97) | 2.55 (0.75, 8.67) | 3.00 (0.36, 24.70) | 1.09 (0.40, 2.96) |
| 40-49 | 2.03 (0.36, 11.19) | **4.36 (1.21, 15.61) \*** | 5.24 (0.58, 47.16) | 1.49 (0.48, 4.61) |
| 50+ | **8.67 (1.37, 54.56)\*** | 1.68 (0.17, 16.36) | **21.66 (2.03, 230.75)\*** | 0.92 (0.10, 8.00) |
| **GENDER** |  |  |  |  |
| Female | Ref | Ref | Ref | Ref |
| Male | 0.57 (0.17, 1.91) | 0.68 (0.30, 1.52) | 0.43 (0.16, 1.13) | 1.03 (0.35, 3.04) |
| **YEAR** |  |  |  |  |
| 2020 | Ref | Ref | Ref | Ref |
| 2021 | **0.26 (0.07, 0.93)\*** | 0.57 (0.25, 1.28) | 0.89 (0.35, 2.27) | 1.10 (0.44, 2.74) |
| 2022 | 0.54 (0.18, 1.55) | **2.90 (1.58, 5.34)\*** | **2.35 (1.03, 5.35) \*** | **3.99 (1.85, 8.60)\*** |
| 2023 | 0.87 (0.36, 2.11) | 0.91 (0.43, 1.96) | 0.40 (0.11, 1.36) | 1.98 (0.84, 4.67) |

aOR – Adjusted Odds Ratio, CI – Confidence Interval, Significant (p<0.05)

**Table 5b:**
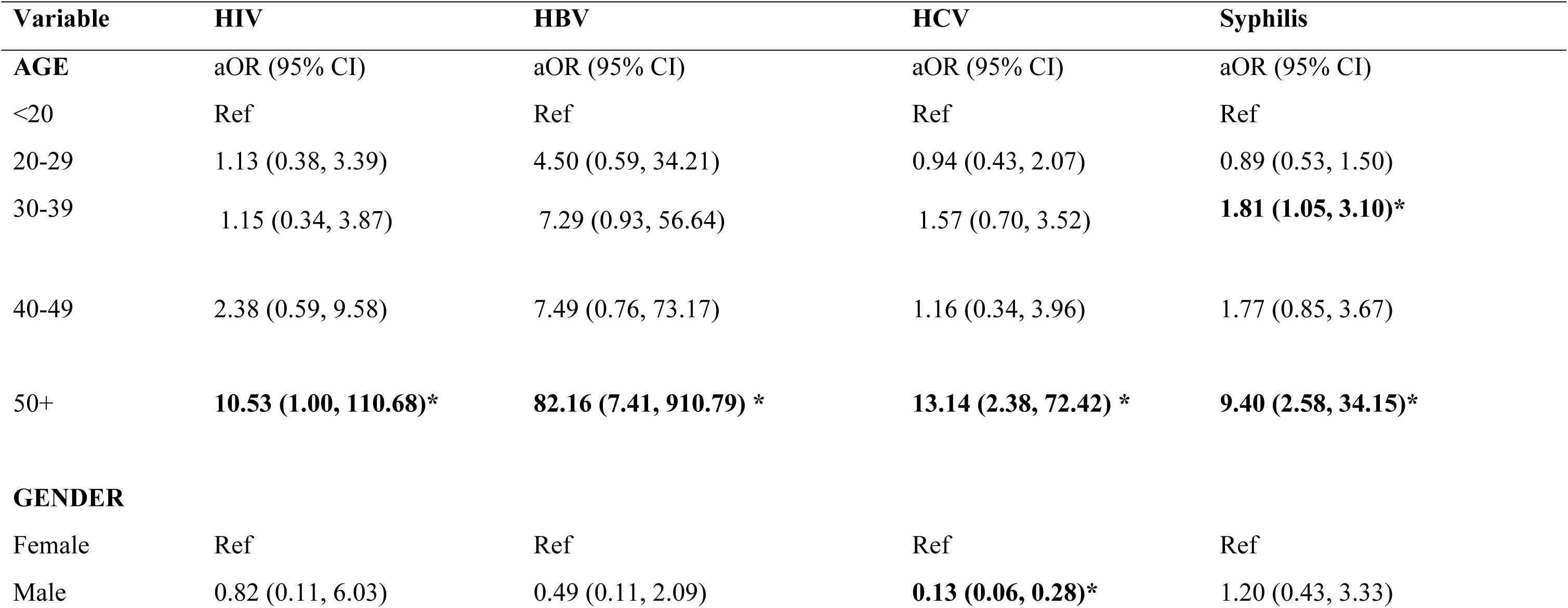
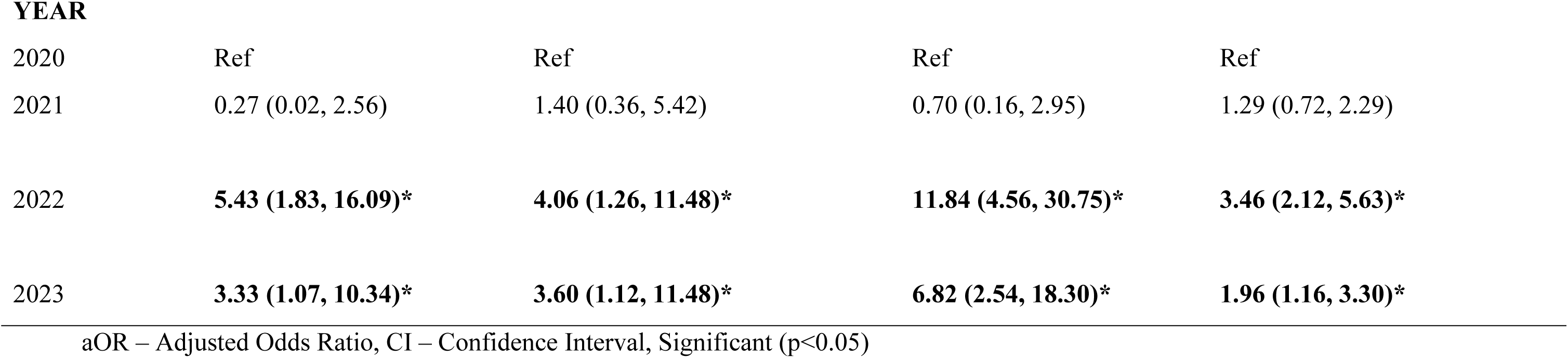
Multivariable logistic regression analysis of factors associated with transfusion-transmissible infections among Hohoe blood donors.

## Discussion

This study assessed the seroprevalence and temporal trends of transfusion-transmissible infections (TTIs) among 6,147 blood donors in Ho and Hohoe over a four-year period. The overall TTI prevalence was low with syphilis (3.6%) as the most common infection, followed by HBV (1.8%), HCV (1.8%), and HIV (1.0%). This pattern suggests improved blood safety compared with earlier Ghanaian reports but indicates persistent residual transfusion risk, particularly for vulnerable recipients (2,8).

The observed prevalence is substantially lower than earlier multicenter Ghanaian estimates reporting overall TTI burdens up to 21% and Hohoe-specific historical prevalence exceeding 16% (2,5). Similar declining trends have been reported in other Ghanaian settings, including Akatsi South (≈8%), suggesting improved donor screening and selection practices across the Volta Region (6). The reduction likely reflects strengthened pre-donation screening, improved laboratory diagnostics, and stricter donor eligibility criteria implemented over recent years (9,10).

Regionally, findings align with variable, but generally higher TTI burdens have been reported across sub-Saharan Africa. In Nigeria, pooled prevalence estimates are approximately 10%, with HBV typically dominant (11). In Gabon, HBV (6%), HCV (4%), HIV (3%), and syphilis (3%) remain higher than in the present study (12). Similarly, Uganda reports an overall prevalence of about 8.7%, with syphilis frequently the leading infection(13). Southern African pooled estimates remain lower (∼2%) but exhibit marked heterogeneity across settings (14).

Globally, high-income countries report TTI prevalence below 1%, largely due to universal voluntary donation systems and advanced screening technologies such as nucleic acid testing (NAT), which significantly reduce window-period infections (15,16). In contrast, many low- and middle-income countries continue to rely on rapid diagnostic tests, which are less sensitive, potentially underestimating viral infections or missing early-stage disease (17).

The predominance of syphilis in this study contrasts with many West African settings where HBV is often the leading TTI but aligns with findings from parts of Ghana, Uganda, and Ethiopia where syphilis is increasingly prominent among donors (13,18). Persistent syphilis transmission may reflect ongoing gaps in sexual health education, untreated community reservoirs, and limited screening outside blood donation contexts (19,20).

Additionally, differences in test sensitivity may contribute to relative pattern shifts. Treponemal rapid tests, while widely used, may yield variability in detection compared with ELISA-based or confirmatory assays, potentially influencing observed prevalence distributions (17). The persistence of syphilis as the leading infection underscores the need for strengthened community-level prevention strategies and improved diagnostic algorithms.

A notable finding is the peak in all TTIs in 2022, followed by a decline in 2023. This pattern is consistent with broader African and global reports documenting post-COVID-19 disruptions in blood safety systems (21,22). During the pandemic, reduced voluntary donations, increased reliance on replacement donors, and strained laboratory capacity likely contributed to temporary increases in TTI prevalence (23,24).

Similar fluctuations have been reported in Ethiopia, Somalia, Cameroon, and Brazil, where health system shocks altered donor recruitment dynamics and screening efficiency (25–27). The subsequent decline in 2023 likely reflects restoration of routine services, improved donor recruitment strategies, and reactivation of public health interventions (10,5). These findings highlight the vulnerability of blood safety systems to external shocks and the importance of resilient haemovigilance frameworks.

Age was a consistent predictor of TTI positivity, with older donors showing higher risk, particularly for HBV and HCV. This is consistent with evidence from multiple African and Asian studies, where cumulative exposure increases infection probability over time (27,18,30). However, some studies also report higher HBV/HCV prevalence in younger donors in high-risk recruitment systems, highlighting context-specific variation (5,16).

Gender differences were more limited but showed a significant association between male sex and HCV positivity in Hohoe. This aligns with studies from Ethiopia and Angola, where males often exhibit higher HBV/HCV prevalence due to greater representation among donors and higher exposure risk behaviors (18,30). Conversely, some settings report higher female seroprevalence due to antenatal screening detection bias, illustrating heterogeneous gender patterns across regions (11,33). Donation year was also a significant predictor, reflecting broader system-level influences such as pandemic disruptions, policy changes, and evolving screening technologies (21,5). Such temporal variability highlights the importance of continuous surveillance in interpreting TTI trends.

A key structural determinant of TTI risk is reliance on replacement or family donors, who consistently demonstrate higher infection rates than voluntary non-remunerated donors (10,33). This remains a major challenge in many African blood systems and likely contributes to persistent infections despite overall declining trends. Socio-demographic factors such as younger age distribution, male dominance, and lower repeat donation rates further exacerbate residual risk (5,6)

### Conclusion

This study determined a TTI seroprevalence of 8.1% among 6,147 blood donors at two blood banks in Ghana’s Volta Region (2020–2023), which is substantially lower than the national multicentered benchmark of 21.0% and prior Hohoe rates exceeding 16.1% (2,34). Seroprevalence peaked in 2022 across all pathogens before declining in 2023. Older age was consistently associated with seropositivity, and site-specific burdens of syphilis (3.6%) and HBV/HCV (1.8% each) were identified. The predominance of young male donors and the contribution of replacement donors emerged as key vulnerabilities, particularly during pandemic-related disruptions.

These findings affirm that sustained implementation of national transfusion safety protocols has strengthened the blood supply. However, persistent reservoirs of TTIs necessitate targeted actions: expanding voluntary non-remunerated donor recruitment, integrating point-of-care syphilis testing, adding anti-HBc screening to detect occult HBV infections, and enhancing female and older-donor recruitment. Future prospective multicenter studies incorporating molecular assays and behavioral risk stratification will be essential to further reduce residual risks and ensure Ghana’s blood system meets universal safety standards.

## Strength and Limitations

### Strengths

This study included all eligible blood donor records over a four-year period, resulting in a large sample size that improves the reliability of the findings. The inclusion of data from two major blood banks in the Volta Region also allowed for comparisons between sites and provides a broader view of transfusion-transmissible infections (TTIs) in the area.

The use of multivariable analysis made it possible to identify key factors associated with TTIs, particularly age and year of donation, while accounting for potential confounding. In addition, the assessment of trends over time provided useful insight into how the prevalence of infections changed across the study period. The use of routinely collected data based on standard screening procedures also makes the findings relevant to real-world blood safety practices.

### Limitations

The retrospective design limits the ability to establish causal relationships and introduces the possibility of unmeasured confounding factors, such as behavioral risks and donor type, including voluntary or replacement donors. This may have influenced the estimated prevalence of transfusion-transmissible infections, as family or replacement donors may conceal high-risk behaviours.

In addition, the study relied on secondary data obtained from routine screening records. As such, the analysis was dependent on the testing methods originally used at the facilities, and it was not possible to verify or standardize these procedures. This may have led to an underestimation of the true prevalence, particularly if infections occurring during the window period were not detected.

The study population was also skewed, with a predominance of male donors and a relatively young age distribution, while females and older individuals were underrepresented. This limits the generalizability of the findings and reduces the statistical power for subgroup analyses by age and gender, a pattern that is consistent with other studies in the region.

## ACKNOWLEDGEMENT

The authors gratefully acknowledge the managements and staff of Hohoe and Ho hospitals and their blood banks for permission to conduct the study and use the data. We also thank the blood bank personnel, laboratory staff, and all individuals who provided technical or administrative support. Special appreciation goes to the blood donors whose records formed the basis of this work.

## Author contribution

AS, TC, AL, EKH and AZD conceptualized and designed the study. AS, and TAY performed the data analysis. AS, TC, AL, DAK, SBB, and KF were responsible for data collection, while AZD supervised the data collection process. AS led the manuscript writing. TAY, EKH, and AZD critically reviewed the manuscript. All authors read and approved the final version of the manuscript.

## Declaration

### Clinical trial number

Not applicable

### Data availability

The data underlying the results presented in this study are available from the corresponding author upon reasonable request.

### Ethics statement

Ethical approval for this study was obtained from the University of Health and Allied Sciences Research Ethics Committee (UHAS-REC), Ghana (Approval number: UHAS-REC B.10 [52]24-25). Permission to access the data was also obtained from the respective health facilities.

The requirement for informed consent was waived by the ethics committee because the study involved secondary analysis of anonymized data with no direct participant contact. All data were handled confidentially and without personal identifiers.

This study was conducted in accordance with the principles of the Declaration of Helsinki.

